# Adapting French COVID-19 vaccination campaign duration to variant dissemination

**DOI:** 10.1101/2021.03.17.21253739

**Authors:** Simon Pageaud, Nicolas Ponthus, Romain Gauchon, Catherine Pothier, Christophe Rigotti, Anne Eyraud-Loisel, Jean-Pierre Bertoglio, Alexis Bienvenüe, François Gueyffier, Philippe Vanhems, Nicolas Leboisne, Jean Iwaz, Stéphane Loisel, Pascal Roy

**Affiliations:** Univ Lyon, Université Claude Bernard Lyon 1, CNRS, Laboratoire de Biométrie et Biologie Evolutive, UMR5558, F-69621, Villeurbanne, France; Pôle Santé Publique, Hospices Civils de Lyon, F-69002, Lyon, France; Univ Lyon, Ecole Centrale de Lyon, ENTPE, CNRS, Laboratoire de Tribologie et Dynamique des Systèmes LTDS, UMR5513, F-69134, Ecully, France; Univ Lyon, Université Claude Bernard Lyon 1, Laboratoire de Sciences Actuarielle et Financière LSAF, ISFA, F-69007, Lyon, France; Univ Lyon, INSA-Lyon, CNRS, Laboratoire d’InfoRmatique en Image et Systèmes d’information LIRIS, UMR5205, F-69621, Villeurbanne, France; INRIA, Grenoble-Rhône-Alpes Research Centre, F-38330, Montbonnot, France; Univ Lyon, Ecole Centrale de Lyon, CNRS, Laboratoire de Mécanique des Fluides et d’Acoustique LMFA, UMR5509, F-69130, Ecully, France; Service Hygiène, Epidémiologie, Infectiovigilance et Prévention, Centre Hospitalier Edouard Herriot, Hospices Civils de Lyon, Lyon, France; Public Health, Epidemiology and Eco-Evolution of Infectious Diseases, CIRI, Centre International de Recherche en Infectiologie, Univ Lyon, Inserm, U1111, Université Claude Bernard Lyon 1, CNRS, UMR5308, ENS de Lyon, F-69007, Lyon, France; Service de Biostatistique-Bioinformatique, Pôle Santé Publique, Hospices Civils de Lyon, F-69002, Lyon, France

## Abstract

**Background:** The outbreak of SARS-CoV-2 virus has caused a major international health crisis with serious consequences in terms of public health and economy. In France, two lockdown periods were decided in 2020 to avoid the saturation of intensive care units (ICU) and an increase in mortality. The rapid dissemination of variant SARS-CoV-2 VOC 202012/01 has strongly influenced the course of the epidemic. Vaccines have been rapidly developed. Their efficacy against the severe forms of the disease has been established, and their efficacy against disease transmission is under evaluation. The aim of this paper is to compare the efficacy of several vaccination strategies in the presence of variants in controlling the COVID-19 epidemic through population immunity.

**Methods:** An agent-based model was designed to simulate with different scenarios the evolution of COVID-19 pandemic in France over 2021 and 2022. The simulations were carried out ignoring the occurrence of variants then taking into account their diffusion over time. The expected effects of three Non-Pharmaceutical Interventions (Relaxed-NPI, Intensive-NPI, and Extended-NPI) to limit the epidemic extension were compared. The expected efficacy of vaccines were the values recently estimated in preventing severe forms of the disease (75% and 94%) for the current used vaccines in France (Pfizer-BioNTech and Moderna since January 11, 2021, and AstraZeneca since February 2, 2021). All vaccination campaigns reproduced an advanced age-based priority advised by the Haute Autorité de Santé. Putative reductions of virus transmission were fixed at 0, 50, 75 and 90%. The effects of four vaccination campaign durations (6-month, 12-month, 18-month and 24-month) were compared.

**Results:** In the absence of vaccination, the presence of variants led to reject the Relaxed-NPI because of a high expected number of deaths (170 to 210 thousands) and the significant overload of ICUs from which 35 thousand patients would be deprived. In comparison with the situation without vaccination, the number of deaths was divided by 7 without ICU saturation with a 6-month vaccination campaign. A 12-month campaign would divide the number of death by 3 with Intensive-NPI and by 6 with Extended-NPI (the latter being necessary to avoid ICU saturation). With 18-month and 24-month vaccination campaigns without Extended-NPI, the number of deaths and ICU admissions would explode.

**Conclusion:** Among the four compared strategies the 6-month vaccination campaign seems to be the best response to changes in the dynamics of the epidemic due to the variants. The race against the COVID-19 epidemic is a race of vaccination strategy. Any further vaccination delay would increase the need of strengthened measures such as Extended-NPI to limit the number of deaths and avoid ICU saturation.

## 1 INTRODUCTION

The outbreak of SARS-CoV-2 virus is causing major national, European, and international health crises, with serious consequences in terms of public health and economy. The COVID-19 epidemic started in China in late autumn 2019 and, on December 31 [1], the WHO China Office [2] received reports on severe pneumonia cases in the city of Wuhan (Hubei Province). On February 14, 2021, more than 100 million cases and 2.4 million officially recognized deaths were recorded worldwide of which 34 million cases and 760 thousand deaths occurred in Europe and 3.4 million cases and 81.6 thousand deaths occurred in France. Europe has been one of the most severely hit continents.

In France, during 2020, two lockdown periods were decided to avoid the saturation of intensive care units (ICU) that would have led to an increase in mortality. The first period (March 17 to May 11, 2020) was decided to limit the magnitude of the first wave. The corresponding maximum 7-day moving average of hospital deaths was 532 on April 8, 2020. Less than 30 daily hospital deaths were observed between June 6 and September 16, defining a plateau. Daily values of 222 and 276 were observed at the beginning (October 30, 2020) and the end (December 15, 2020) of the second lockdown, with a maximum 7-day moving average of hospital deaths of 419 on November 19, 2020. Nevertheless, the resulting epidemic situation was less favorable after the second lockdown; there was a residual plateau with 240 to 340 daily deaths.

On December 14, 2020, the variant SARS-CoV-2 VOC 202012/01 (B.1.1.7) was identified in United Kingdom [3]. The first case with this variant appeared in France on December 2020. It was deemed responsible of 3.3% of all new cases on January 8, 2021 [4], 14% on January 27 [5], and 35% on February 16. By the end of February 2021, this percentage level had reached 50%. Preliminary studies suggest that SARS-CoV-2 VOC 202012/01 increased transmissibility (infectiousness) but not disease severity.

Vaccines have been rapidly developed. Their efficacy against the severe forms of the disease was estimated between 75% [6] and 94% [7, 8]. The efficacy of these vaccines on the disease transmission is still under evaluation. The objective of the current massive vaccination strategy in France is to achieve the extinction of the epidemic through herd immunity resulting from an extended vaccination coverage.

An important public health issue is to optimize the population vaccination schedule and ensure that necessary protective measures are adapted to the epidemic situation and its evolution. The dynamics of the epidemic is continuously assessed through changes in hospital admissions, intensive care units (ICU) occupancy rate, and cumulative number of deaths associated with COVID-19. In France, protective measures have been constantly adapted to the dynamics of the epidemic. These measures are monitored using the value of the effective reproductive number and the weekly incidence rate.

The effectiveness of a vaccination campaign depends strongly on the availability of vaccine vials and on their distribution strategy. A campaign should also adapt to the evolution of the epidemic, especially in the presence of variants. This study aims to analyze the expected dynamics of the COVID-19 epidemic after applying protective measures and taking into account the increasing proportion of more infectious variants (e.g. SARS-CoV-2 VOC 202012/01) and several vaccination strategies.

## 2 MATERIALS AND METHODS

### 2.1 Compartmental model

The model is a compartmental model, stratified by age groups and adapted from the model described in [9]. A graphical view of the model can be found in [10]. The model compartments are the following:

- *S* (Susceptible): individuals who are not infected by the virus;
- The incubation period included two compartments:
  – when infected, an individual goes from *S* to compartment *E* (Exposed) that contains infected individuals who did not develop symptoms yet and are not contagious. The mean length of stay in *E* is (*t*_*i*_ *− t*_*p*_) where *t*_*i*_ is the incubation period and *t*_*p*_ the duration of the prodromal state (see next item);
  – when an individual from *E* starts to be contagious, it is transferred to compartment *I*_*p*_ (Prodromal phase which is the short phase that follows contamination without pathological symptoms but with possible non-specific prodromes) that contains individuals still in the incubation period already contagious but without apparent symptoms. After a stay of average duration *t*_*p*_, the individuals enter one of the four compartments *A, I*_*ps*_, *I*_*ms*_, and *I*_*ss*_ defined hereafter.
- *A* (Asymptomatic): individuals who completed the incubation period, became infectious, but have no disease symptoms (with probability *p*_*A*_). The mean length of stay in *A* is *t*_*s*_;
- The symptomatic infectious period includes three compartments for individuals developing symptoms (with probability 1 *− p*_*A*_):
  – *I*_*ps*_ (Paucisymptomatic disease): individuals with weak disease symptoms;
  – *I*_*ms*_ (Medium symptoms): individuals with disease symptoms (e.g. fever or cough) who will not require hospitalization. The average durations in states *I*_*ps*_ and *I*_*ms*_ are the same as the duration in state *A*;
  – *I*_*ss*_ (Severe symptoms): individuals severely infected, who will require hospitalization. They stay in *I*_*ss*_ before being hospitalized. The mean length of stay in *I*_*ss*_ is *t*_*bh*_.
  Once symptomatic, the probabilities to enter *I*_*ps*_, *I*_*ms*_, or *I*_*ss*_ are respectively *p_Ips_, p_Ims_*, and *p*_*H*_ (probabilities summing to one).
- The hospitalization period includes two compartments:
  – *ICU*: hospitalized individuals who during part of their stay will require an ICU bed;
  – *H* (Hospitalized): hospitalized individuals who will not require ICU care during their stay.
  Individuals coming from compartment *I*_*ss*_ are admitted to *ICU* with probability *p*_*ICU*_ and to *H* with probability 1 *− p*_*ICU*_. The mean length of stay in *ICU* (resp. *H*) is *t*_*icu*_ (resp. *t*_*h*_).
- In this model, there are two absorbing compartments:
  – *D* (Deceased at hospital): probability to enter *D* after *ICU* (resp. *H*) is *p*_*D*|*ICU*_ (resp. *p*_*D*|*H*_);
  – *R* (Removed): individuals who recovered (from any compartment).

The dynamics can be expressed according to the following system of ordinary differential equations where *α* denotes the incidence hazard rate:

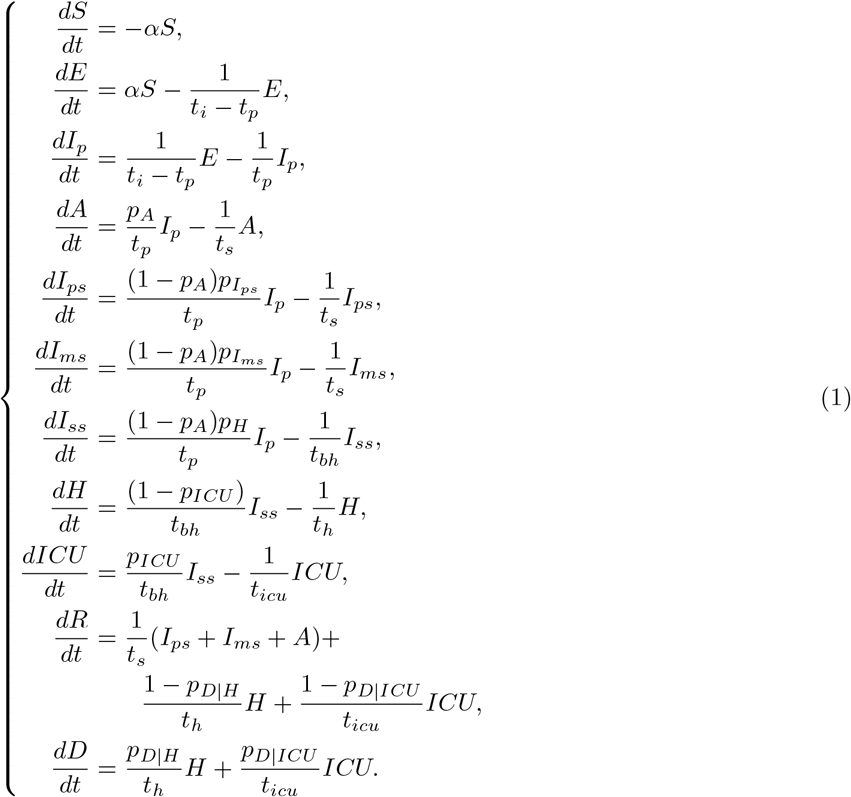

Here, each compartment was split into 17 age groups, as in [10], whereas only three age groups were considered in [9]. The incidence hazard rate *α* could vary over time and was different between each age groups (as defined in the next section). Parameter values are given in tables 1 and 2.

**Table 1:** Simulation parameters, sources see [10].

| age | 0-4 | 5-9 | 10-14 | 15-19 | 20-24 | 25-29 | 30-34 | 35-39 | 40-44 | 45-49 |
| --- | --- | --- | --- | --- | --- | --- | --- | --- | --- | --- |
| $p_{I_{ps}}$ | 0.25 | 0.25 | 0.249 | 0.249 | 0.247 | 0.247 | 0.242 | 0.242 | 0.238 | 0.238 |
| $p_{I_{ms}}$ | 0.749 | 0.749 | 0.748 | 0.748 | 0.741 | 0.721 | 0.726 | 0.726 | 0.713 | 0.713 |
| $p_{I_{ss}}$ | 0.001 | 0.001 | 0.003 | 0.003 | 0.012 | 0.012 | 0.032 | 0.032 | 0.049 | 0.049 |
| $t_h$ | 8.1 | 8.1 | 8.1 | 8.1 | 8.1 | 8.1 | 7.4 | 7.4 | 8 | 8 |
| $t_{icu}$ | 19.8 | 19.8 | 19.8 | 19.8 | 19.8 | 19.8 | 20.7 | 20.7 | 20.5 | 20.5 |
| $p_{ICU}$ | 0.285 | 0.285 | 0.285 | 0.285 | 0.285 | 0.285 | 0.323 | 0.323 | 0.343 | 0.343 |
| $p_{D\vert H}$ | 0 | 0 | 0 | 0 | 0 | 0 | 0.023 | 0.023 | 0.058 | 0.058 |
| $p_{D\vert ICU}$ | 0 | 0 | 0 | 0 | 0 | 0 | 0.2 | 0.2 | 0.31 | 0.31 |

| age | 50-54 | 55-59 | 60-64 | 65-69 | 70-74 | 75-79 | 80+ |
| --- | --- | --- | --- | --- | --- | --- | --- |
| $p_{I_{ps}}$ | 0.225 | 0.225 | 0.209 | 0.209 | 0.189 | 0.189 | 0.182 |
| $p_{I_{ms}}$ | 0.674 | 0.674 | 0.626 | 0.626 | 0.568 | 0.568 | 0.545 |
| $p_{I_{ss}}$ | 0.102 | 0.102 | 0.166 | 0.166 | 0.243 | 0.243 | 0.273 |
| $t_h$ | 9.4 | 9.4 | 11.4 | 11.4 | 15.9 | 15.9 | 26.6 |
| $t_{icu}$ | 24.5 | 24.5 | 17.6 | 17.6 | 23.4 | 23.4 | 19.2 |
| $p_{ICU}$ | 0.408 | 0.408 | 0.36 | 0.36 | 0.282 | 0.282 | 0.05 |
| $p_{D\vert H}$ | 0.081 | 0.081 | 0.147 | 0.147 | 0.227 | 0.227 | 0.434 |
| $p_{D\vert ICU}$ | 0.25 | 0.25 | 0.47 | 0.47 | 0.54 | 0.54 | 0.47 |

**Table 2:** Simulation parameters.

| Parameter | Sources historical strain | Historical strain | New variants |
| --- | --- | --- | --- |
| $t_i$ | see [10] | 5.1 | 5.1 |
| $t_p$ | see [10] | 1.5 | 1.5 |
| $p_A$ | [9, 20] | 0.20 or 0.35 | 0.20 or 0.35 |
| $i_A$ | [21] | 0.55 | $1.7 \times 0.55 = 0.935$ |
| $i_{Ip}$ | [21] | 0.55 | $1.7 \times 0.55 = 0.935$ |
| $i_{Ips}$ | [21] | 0.55 | $1.7 \times 0.55 = 0.935$ |
| $i_{Im_s}$ | [21] | 1 | 1.7 |
| $i_{Iss}$ | [21] | 1 | 1.7 |
| $\beta_1$ | calibration | 0.05052 or 0.05331 | 0.05052 or 0.05331 |
| $t_s$ | see [10] | 7 | 7 |
| $t_{bh}$ | see [10] | 3 | 3 |

### 2.2 Modeling the disease contagiousness

For a given age group *i*, at a given time *t*, the incidence hazard rate 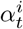 was calculated as follows:

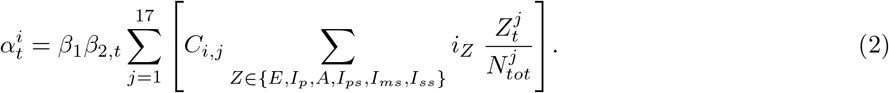

where:

- *C* is the contact matrix where an element *C*_*i,j*_ is the average number of individuals from age group *j* encountered per day by a single individual from age group *i*;
- 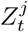 is the number of individuals of age group *j* in compartment *Z* at time *t*;
- 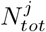 is the total number of individuals from age group *j* (who had contracted the disease or not);
- *i*_*Z*_ is the relative infectiousness of individuals in compartment *Z*. For the historical strain, it is set to 1 in case of medium or severe symptoms; it ranges from 0 to 1 for the other compartments;
- *β*_1_ is the daily incidence hazard rate. In can be interpreted as a hazard rate in case of one daily contact with a medium- or severe-symptom case infected by the historical strain;
- The relative hazard *β*_2,*t*_ estimates the effect of Non-Pharmaceutical Interventions (NPI), also named barrier measures, such as mask wearing, social distancing, lockdown, and auto-isolation after infectious contacts. The underlying hypothesis is that NPI effects are similar in all compartments and age groups, with a multiplicative effect on the hazard (without multiplicative interaction). *β*_2,*t*_ can be interpreted as a coefficient reducing the mean number of daily contacts and/or the mean duration of contact, and/or their infectiousness. The introduction of barrier measures leads to 0 *≤ β*_2,*t*_ *≤* 1, by ignoring here an improbable exacerbation of risk behaviors (*β*_2,*t*_ *>* 1) once the epidemic is declared.

### 2.3 Reproductive numbers

At an epidemic start, almost all individuals are susceptible and no protective measure are already taken. The effective reproductive number *R*_*e*_(*t*) is then at its maximum and corresponds to the basic reproductive number *R*_0_ which characterizes the virus infectiousness and the initial environmental conditions of viral spread (i.e. before the initiation of the NPI). The reproductive numbers *R*_0_ and *R*_*e*_(*t*) give an indication of the epidemic dynamics: the highest they are, the faster the epidemic spreads. The effective number *R*_*e*_(*t*) decreases with time, following the decreased proportion of susceptible individuals combined with the introduction of NPI and is defined by

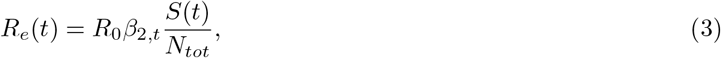

where *N*_*tot*_ is the total size of the population.

To compute *R*_0_ as defined by Diekmann *et al*. [11], it is necessary to introduce the concept of next generation matrix. In the following, the approach and notational conventions of Van den Driessche and Watmough [12] are used. It is assumed that there are *m* different compartments of infected individuals and *n* different age groups. Thus, there is a total of *nm* groups of infected (but not necessarily infectious) individuals. Let *x*(*t*) = (*x*_1_(*t*), …, *x*_*nm*_(*t*)), with quantity *x*_*i*_(*t*), 1 ≤ *i* ≤ nm, denote the number of individuals in the *i*th group of infected individuals at time *t*. Let *ℱ*_*i*_(*x*(*t*)) be the rate of new infected individuals in the *i*th group of infected individuals at time *t*. Let 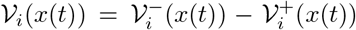 where 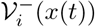 is the rate of departures from group *i*, and 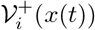 is the rate of arrivals in group *i* by all other means than new infections. Let 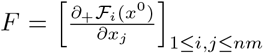 and 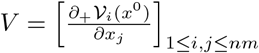 where *x*^0^ is a disease free equilibrium and *∂*_+_ denotes right partial derivatives. The next generation matrix is then defined as

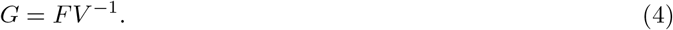

As shown in [12], 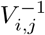 can be seen as the average time an individual introduced in the *j*th group of infected individuals will spend in the *i*th group of infected individuals during the remaining lifetime. Thus, intuitively, an entry (*i, j*) of the next generation matrix *FV* ^*−*1^ is the expected number of new cases in group *i* due to individuals introduced in group *j. R*_0_ is then defined as the spectral radius of *G*. It is computed before initiation of the NPI (i.e., with *β*_2,*t*_ = 1), then, as shown in [10], the expression of *R*_0_ is of the form *R*_0_ = *kβ*_1_ with *k* depending on the other parameters. In the following, using Equation (3), this provides the relationship to calculate *β*_2,*t*_ from *R*_*e*_ after *β*_1_ has been estimated.

### 2.4 Agent-based simulation of the pandemic

The model was implemented as a stochastic Agent-Based Model (ABM). A discrete time step of one day was used for the simulations assuming that the derivatives in the equation system (1) were constant functions over this interval.

#### 2.4.1 Parameters

The agent states corresponded to the compartments, and the probability to exit from a state was obtained from the mean length of stay in the associated compartment. When more than one destination state were possible, the transition is selected at random using the corresponding probabilities (i.e., *p*_*A*_, *p*_*Ips*_, *p*_*Ims*_*, p*_*H*_, *p*_*ICU*_, *p*_*D*|*H*_ and *p*_*D*|*ICU*_).

The age distribution of the population came from INSEE (French Institut National de la Statistique et des Etudes Economiques) studies; it considered 16 five-year age groups from 0 to 79 years plus an extra age-group with individuals aged 80 or older. The contact matrix was available for France on the basis of these age ranges [13]. The number of daily contacts per individual was set at the beginning of the simulation and was drawn from a Poisson distribution with a parameter set to the corresponding element of the contact matrix. At each time step, the daily contacts between infectious and susceptible agents were drawn uniformly at random. For each such contact, the probability of transmission was determined by *β*_1_, *β*_2,*t*_ and the value of the relative infectiousness coefficient associated with the compartment of the infectious agent.

The proportion of asymptomatic forms was set at 20% and 35% (Table 2). The proportions of various symptomatic forms, paucysymptomatic, medium symptom and severe symptom were specified as depending on age (Table 1). The relative infectiousness of prodromal, asymptomatic, and paucysymptomatic forms of the historical strain was set to 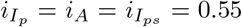 medium and severe symptom forms were taken as references with 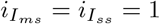 (Table 2).

Parameter *β*_1_ was estimated using the same method as [10], by fitting hospitalization data before and during the first lockdown in France. The maximum likelihood estimation led to *β*_1_ = 0.05052 when the asymptomatic rate was 20% and to *β*_1_ = 0.05331 when the asymptomatic rate was 35%. On January 1, 2021, the beginning of the simulations, the estimated proportion of removed individuals was 13% [14] and the proportion of variant considered negligible [15].

#### 2.4.2 Scenarios

Three different scenarios were considered: Relaxed-NPI, Intensive-NPI, and Extended-NPI. These scenarios took into account a mix of several protective measures taken to reach a specific effective reproduction number *R*_*e*_. The three typical groups of protective measures were designed from the French restrictions observed during the past year: i) *Intensive protective measures* consisted in a combination of strict protective measures, such as overnight curfew, social distancing, mask-wearing, increased distancing at work, closing of bars, restaurants and leisure facilities, etc.; ii) *Extended protective measures* consisted in very strict sanitary measures enhanced compared to the intensive ones, such as weekend or full lockdown, as well as closing of schools, universities, and shopping facilities; iii) *Moderate protective measures* consisted in relaxing the intensive measures (such as restaurant and leisure facility reopening) but maintaining some protective measures such as mask wearing and social distancing. The Intensive-NPI included constant intensive protective measures. The Extended-NPI included constant extended protective measures. The Relaxed-NPI associated systematic alternation of 45-days periods with Intensive protective measures and Moderate protective measures.

These three scenarios that corresponded to different protective procedures were specified through the value of the relative hazard *β*_2,*t*_, under the hypothesis of a similar multiplicative effect of protective measures whatever the viral strain, without interaction. On January 1, 2021, the estimated effective reproductive number was *R*_*e*_ = 1 [15]. For *R*_*e*_ = 1, the calculated relative hazard at this date was *β*_2,*t*_ = 0.2501. This value of parameter *β*_2,*t*_ was the value associated with the so-called *Intensive protective measures*, in order to match the effective situation of the sanitary protective measures adopted in France on January 1, 2021. The values of parameter *β*_2,*t*_ for the other protective measures were calculated with hypothetical measures leading to different values of *R*_*e*_: *R*_*e*_ = 0.8 (*Extended protective measures, β*_2,*t*_ = 0.2001) and *R*_*e*_ = 1.5 (*Moderate protective measures, β*_2,*t*_ = 0.3752) on January 1, 2021.

The expected effects of the three protective scenarios were compared. Due to the stochasticity of the agent-based simulation, 50 simulations were performed with each set of parameters. Each run involved 322,488 agents (i.e., 0.5% of the population of metropolitan regions of France, of about 64.5 million inhabitants). To obtain estimations for France, simulation results were multiplied by 200. The mean values and standard errors of the mean across the runs were plotted.

#### 2.4.3 New variant influence

The so-called ‘variant strain’ herein corresponds to several variants considered more contagious than the historical strain. The relative infectiousness of the variant strains vs. the historical strain were supposed independent of the compartment. This corresponded to multiplying each *i*_*z*_ by a parameter *β*_3_; the latter parameter was the same whatever the compartment and was characteristic of the relative infectiousness of the variant strain. The value of *β*_3_ fitting with a prediction of a proportion of variants close to the available data on January 8, 2021 (3.3%), January 27, 2021 (14%), and February 18, 2021 (36%) [4, 5, 16] was found to be *β*_3_ = 1.7, with at the beginning of the simulation (January 1, 2021), an intercept of the new variant rounded up to 2% of the population infected with the historical strain.

### 2.4.4 Vaccine effects

The expected effects of vaccination were set at their recently estimated levels of efficacy in preventing severe forms of the disease; that is, 94% [7, 8] and 75% [6], depending on the vaccine. When the vaccine was effective, a vaccinated individual could develop only an asymptomatic form of the disease. Putative reductions of virus transmission were fixed at 0, 50, 75 and 90%, the transmission probabilities being considered independent of the vaccine. An age-based priority for being vaccinated was used. Three vaccination priorities were considered and all individuals of a given age group had to receive their first injection before giving the first injections to the next priority age group. The age priorities considered were: 75 and older, 65 and older, then the rest of the population. In the rest of the population, two vaccination options were compared: vaccinate at all ages vs. vaccinate individuals aged 20 years and over (vaccination is under investigation in the youngest population). The effects of four vaccination campaigns over 6, 12, 18, 24 months were compared. At the end of the vaccination campaign, all individuals eligible to vaccination had to have received their second injection. In all simulations, vaccines with 94% efficacy were attributed to individuals aged 75 or older, whereas vaccines with 75% efficacy were attributed to the rest of the population. To be considered as vaccinated, an individual had to receive two injections of any vaccine with a 21-day interval. In all simulations, vaccine supplies were considered sufficient to follow the study’s vaccination schedules. Furthermore, all vaccines were considered as efficient against the historical strain as against new variants similar to the B.1.1.7 lineage [17].

#### 2.4.5 Outcome criteria

The main outcome was the cumulative number of individuals removed, the cumulative number of deaths at hospital, the daily prevalence in ICU beds and its saturation indicator. The saturation of ICU beds was calculated as the cumulative number of new cases requiring ICU when all beds were already occupied. Three thresholds were considered: 5,000 (the current number of ICU beds), 8,000 (adding beds in intermediate care units), and 12,000 (adding also more beds in conventional care units). The model specified that, on average, people in the intensive care compartment stayed the same time in an intensive care bed or a conventional hospital bed^1^. The cumulative number of individuals removed due to disease was calculated as of December 31, 2022.

## 3 RESULTS

### 3.1 Dynamics of the epidemic with the historical strain without vaccination

Table 3 presents the results obtained by restricting the simulations to the historical strain and for 20% and 35% prevalence of asymptomatic cases, without vaccination. After two years of Relaxed-NPI, the simulated numbers of removed individuals were 20.4 and 20.5 millions and the numbers of deaths were 62.6 and 51.2 thousand respectively. The simulated numbers of ICU admissions that exceeded the 5,000-bed threshold were 2,860 and 1,890, respectively. In this scenario, the cumulative distribution of deaths plateaued after 500 days (Figure 1a). The waves observed for the Relaxed-NPI scenarios show the slightly delayed effects of the systematic alternation of periods of 45 days with intensive protection measures and moderate protection measures (Figure 1b). After an initial decrease, three successive waves were observed at the end of the three first 45-days periods with Moderate protective measures. The second wave exceeded ICU capacities (Figure 1b), whereas the third wave showed a decrease before an extinction of the epidemic.

**Table 3:**
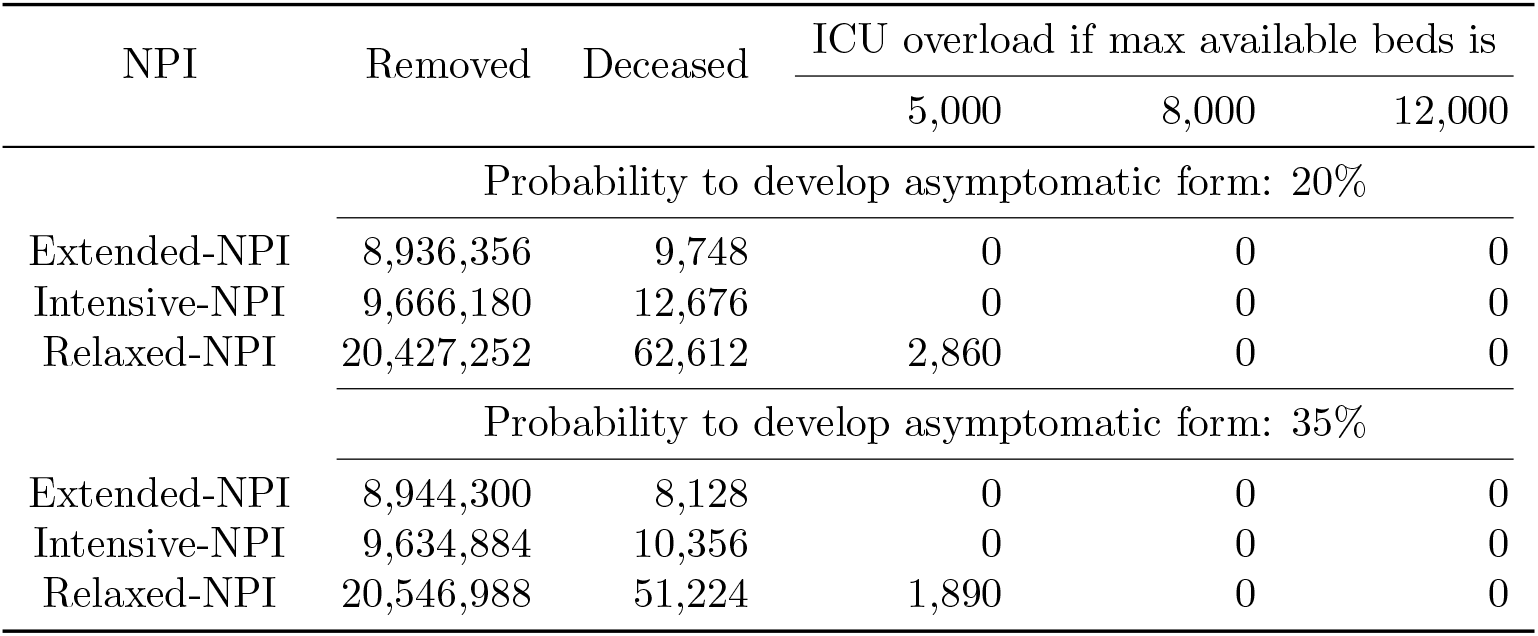
historical strain without vaccination: two-years cumulative numbers of individuals removed and hospital deaths, ICU overload exceeding beds capacity under different restrictions.

**Figure 1:**
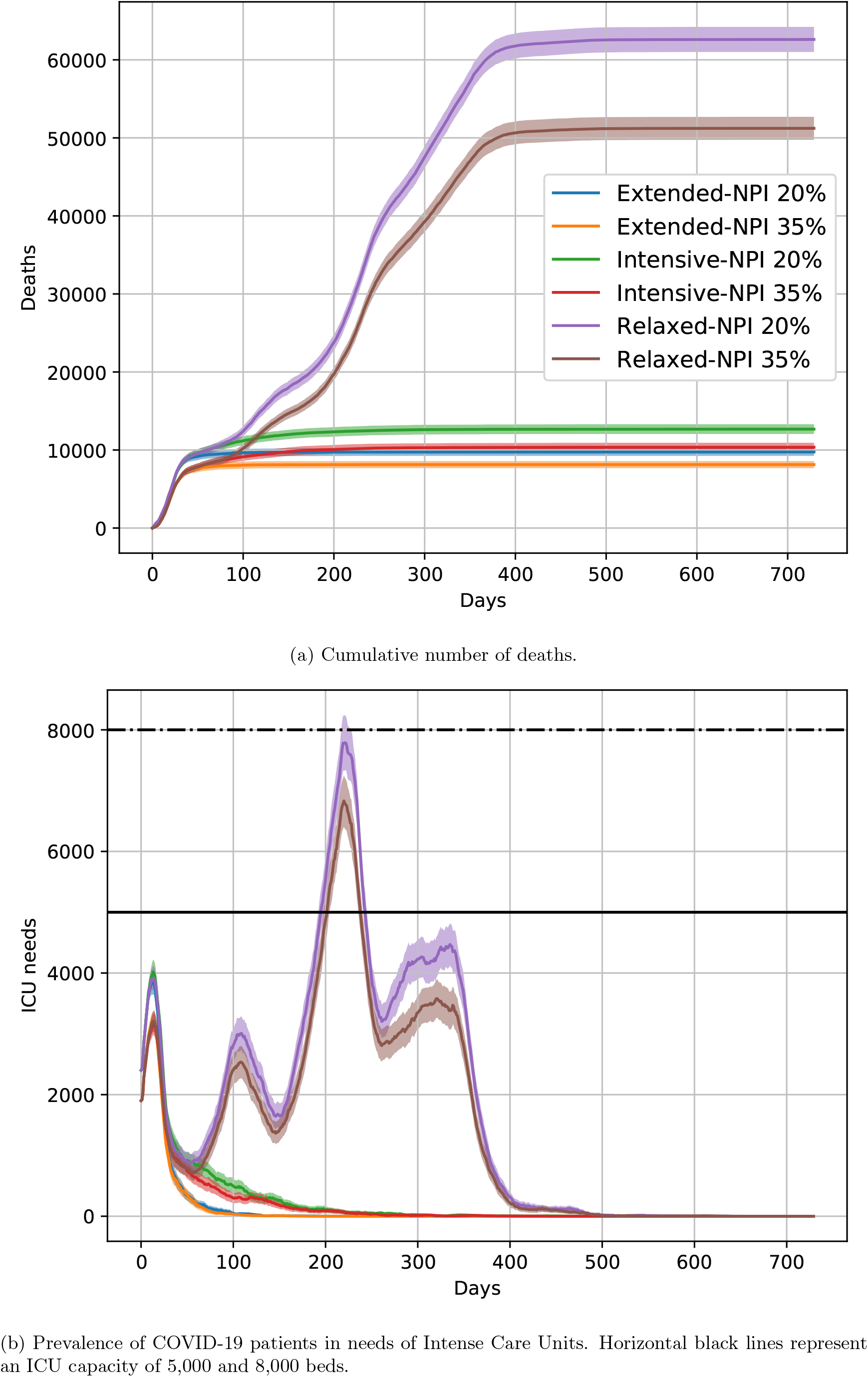
Projection of the COVID-19 pandemic in France for the historical strain, without vaccine, for Extended-NPI, Intensive-NPI and Relaxed-NPI, and for 20% and 35% asymptomatic rates.

After two years of the Intensive-NPI, the simulated numbers of removed individuals was much lower (around 9.6 million) with 12.7 and 10.4 thousand deaths, respectively. The cumulative distribution of deaths plateaued after 250 days (Figure 1a). The ICU capacities were not exceeded. A rapid decrease was observed whatever the prevalence of asymptomatic cases (Figure 1b) leading to the extinction of the epidemic.

After two years of the Extended-NPI, the simulated numbers of removed individuals were around 8.9 million with 9.7 and 8.1 thousand deaths, respectively. The cumulative distribution of deaths plateaued after 100 days (Figure 1a). The ICU capacities were not exceeded. A rapid decrease was observed whatever the prevalence of asymptomatic cases (Figure 1b) leading to the extinction of the epidemic.

### 3.2 Dynamics of the epidemic with the introduction of new variant strains without vaccination

Table 4 presents the results obtained taking into account a competition between the historical strain and the more infectious new variants with 20% and 35% prevalence of asymptomatic cases, without vaccination. Applying the Relaxed-NPI, the simulated numbers of removed individuals were around 40.2 million, with 207.7 and 168.5 thousand deaths. The cumulative distribution of deaths plateaued after 250 days (Figure 2a). The simulated numbers of ICU admissions that exceeded the 5,000-bed threshold were 35.1 and 27.2 thousand respectively. The capacities of the ICU were also saturated beyond 8,000 or 12,000 beds. A huge single wave was observed, resulting from the fusion of the effects of the two first 45-days periods with moderate protective measures, leading to the extinction of the epidemic after this single wave (Figure 2b). Given these simulated results, the Relaxed-NPI was not further retained.

**Table 4:** historical strain and variant development without vaccination: two-years cumulative numbers of individuals removed and hospital deaths, ICU overload exceeding beds capacity under different restrictions.

| NPI | Removed | Deceased | ICU overload if max available beds is |  |  |
| --- | --- | --- | --- | --- | --- |
|  |  |  | 5,000 | 8,000 | 12,000 |
| Probability to develop asymptomatic form: 20% |  |  |  |  |  |
| Extended-NPI | 21,788,476 | 70,176 | 1,914 | 0 | 0 |
| Intensive-NPI | 32,460,452 | 137,556 | 24,326 | 21,326 | 17,326 |
| Relaxed-NPI | 40,223,456 | 207,732 | 35,060 | 32,060 | 28,060 |
| Probability to develop asymptomatic form: 35% |  |  |  |  |  |
| Extended-NPI | 21,818,740 | 56,416 | 672 | 0 | 0 |
| Intensive-NPI | 32,482,356 | 111,332 | 18,940 | 15,940 | 11,940 |
| Relaxed-NPI | 40,250,780 | 168,536 | 27,192 | 24,192 | 20,192 |

**Figure 2:**
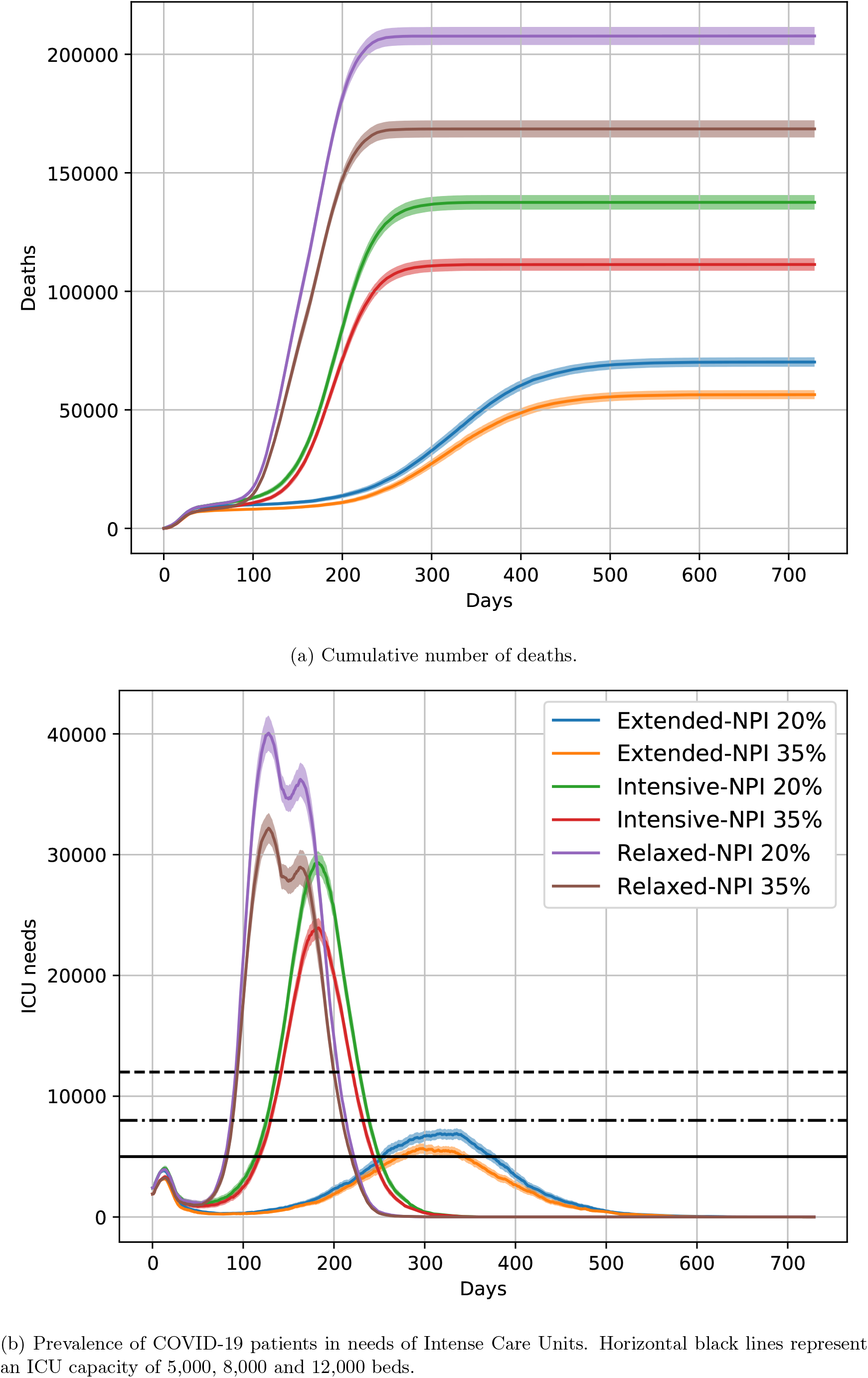
Projection of the COVID-19 pandemic in France with the historical strain and new variants, without vaccine, for Extended-NPI, Intensive-NPI and Relaxed-NPI, and for 20% and 35% asymptomatic rates.

When the Intensive-NPI was applied, the simulated numbers of removed individuals were still important (around 32.5 million) with 137.6 and 111.3 thousand deaths, respectively. The cumulative distribution of deaths plateaued after 300 days (Figure 2a). The simulated saturation of ICU capacities over 5,000 beds concerned 24.3, and 18.9 thousand patients. The capacities of the ICU were saturated beyond 8,000 or 12,000 beds. A huge wave was observed, despite the keep up of the Intensive-NPI, leading to the extinction of the epidemic after a massive contamination of the population (Figure 2b).

When the Extended-NPI were applied, the simulated numbers of removed individuals were 21.8 million with 70.2 and 56.4 thousand deaths, respectively. The cumulative distribution of deaths plateaued after 550 days (Figure 2a). The simulated numbers of ICU admissions that exceeded the 5,000-bed threshold were 1.9 and 0.7 thousand. The capacities of the ICU were never saturated beyond 8,000 beds. Compared to that simulated under Intensive-NPI, the wave observed under Extended-NPI was delayed with a maximum at 300 days and an extinction of the epidemic after 550 days (Figure 2b).

### 3.3 Effects of a 6-month vaccination campaign

Table 5 shows the results obtained with a 6-month vaccination campaign.

**Table 5:** Results of a 6-month vaccination campaign with the historical strain and new variants: two-years cumulative numbers of individuals removed and deaths, ICU overload exceeding beds capacity under different restrictions, different vaccination options and different transmission reduction.

| NPI | Vaccination<br>Option | Transmission<br>Reduction | Removed | Deceased | ICU overload if max available beds is |  |  |
| --- | --- | --- | --- | --- | --- | --- | --- |
|  |  |  |  |  | 5,000 | 8,000 | 12,000 |
| $p_A = 20\%$ | | | | | | | |
| Extended NPI | 20+ | 0% | 11,183,064 | 10,352 | 0 | 0 | 0 |
| Extended-NPI | 20+ | 50% | 9,320,296 | 9,928 | 0 | 0 | 0 |
| Extended-NPI | 20+ | 75% | 9,220,956 | 10,316 | 0 | 0 | 0 |
| Extended-NPI | 20+ | 90% | 9,146,692 | 10,132 | 0 | 0 | 0 |
| Extended-NPI | All | 0% | 9,434,808 | 10,244 | 0 | 0 | 0 |
| Extended-NPI | All | 50% | 9,089,316 | 9,632 | 0 | 0 | 0 |
| Extended-NPI | All | 75% | 9,059,508 | 9,832 | 0 | 0 | 0 |
| Extended-NPI | All | 90% | 9,041,424 | 10,088 | 0 | 0 | 0 |
| Intensive-NPI | 20+ | 0% | 24,716,196 | 18,236 | 0 | 0 | 0 |
| Intensive-NPI | 20+ | 50% | 15,598,584 | 15,300 | 0 | 0 | 0 |
| Intensive-NPI | 20+ | 75% | 13,679,532 | 14,636 | 0 | 0 | 0 |
| Intensive-NPI | 20+ | 90% | 12,793,640 | 14,028 | 0 | 0 | 0 |
| Intensive-NPI | All | 0% | 19,373,548 | 16,100 | 0 | 0 | 0 |
| Intensive-NPI | All | 50% | 10,938,868 | 13,428 | 0 | 0 | 0 |
| Intensive-NPI | All | 75% | 10,316,708 | 12,696 | 0 | 0 | 0 |
| Intensive-NPI | All | 90% | 10,089,244 | 12,792 | 0 | 0 | 0 |
| $p_A = 35\%$ | | | | | | | |
| Extended-NPI | 20+ | 0% | 12,443,196 | 8,572 | 0 | 0 | 0 |
| Extended-NPI | 20+ | 50% | 9,309,228 | 8,260 | 0 | 0 | 0 |
| Extended-NPI | 20+ | 75% | 9,193,656 | 8,100 | 0 | 0 | 0 |
| Extended-NPI | 20+ | 90% | 9,194,984 | 8,312 | 0 | 0 | 0 |
| Extended-NPI | All | 0% | 9,666,408 | 8,424 | 0 | 0 | 0 |
| Extended-NPI | All | 50% | 9,110,852 | 7,812 | 0 | 0 | 0 |
| Extended-NPI | All | 75% | 9,067,128 | 8,348 | 0 | 0 | 0 |
| Extended-NPI | All | 90% | 9,052,396 | 8,084 | 0 | 0 | 0 |
| Intensive-NPI | 20+ | 0% | 26,017,820 | 15,480 | 0 | 0 | 0 |
| Intensive-NPI | 20+ | 50% | 16,085,204 | 12,868 | 0 | 0 | 0 |
| Intensive-NPI | 20+ | 75% | 13,764,324 | 11,892 | 0 | 0 | 0 |
| Intensive-NPI | 20+ | 90% | 13,028,460 | 11,800 | 0 | 0 | 0 |
| Intensive-NPI | All | 0% | 21,608,704 | 13,708 | 0 | 0 | 0 |
| Intensive-NPI | All | 50% | 11,118,796 | 11,208 | 0 | 0 | 0 |
| Intensive-NPI | All | 75% | 10,317,812 | 10,000 | 0 | 0 | 0 |
| Intensive-NPI | All | 90% | 10,123,672 | 10,268 | 0 | 0 | 0 |

Applying the Intensive-NPI with a 20% prevalence of asymptomatic forms, a 90% reduction in virus transmission halved the number of removed and reduced the number of deaths by 20-25% vs. no vaccine effect on virus transmission.

In comparison with vaccination of individuals aged 20 or older, the vaccination of all individuals reduced the number of deaths by 10%.

The ICU resources were never saturated.

With a 6-month vaccination campaign of all individuals without transmission reduction (Table 5) vs. no vaccination and Intensive-NPI (Table 4), the number of deaths was divided by 9 with the Intensive-NPI and by 13 with the Extended-NPI.

### 3.4 Effects of a 12-month vaccination campaign

Table 6 shows the results obtained with a 12-month vaccination campaign.

**Table 6:**
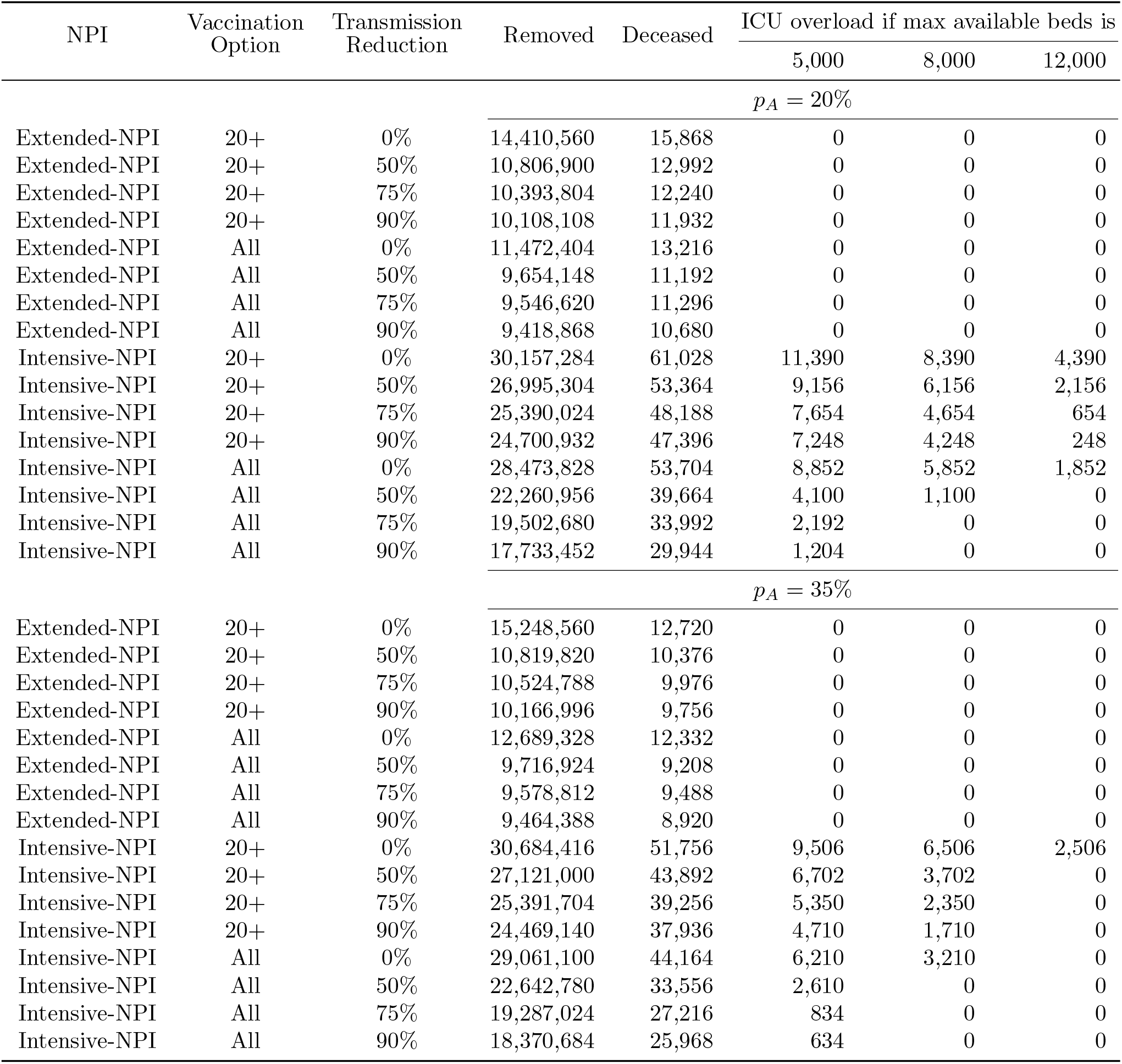
Results of a 12-month vaccination campaign with the historical strain and new variants: two-years cumulative numbers of individuals removed and deaths, ICU overload exceeding beds capacity under different restrictions, different vaccination options and different transmission reduction.

| NPI | Vaccination<br>Option | Transmission<br>Reduction | Removed | Deceased | ICU overload if max available beds is |  |  |
| --- | --- | --- | --- | --- | --- | --- | --- |
|  |  |  |  |  | 5,000 | 8,000 | 12,000 |
| $p_A = 20\%$ | | | | | | | |
| Extended-NPI | 20+ | 0% | 14,410,560 | 15,868 | 0 | 0 | 0 |
| Extended-NPI | 20+ | 50% | 10,806,900 | 12,992 | 0 | 0 | 0 |
| Extended-NPI | 20+ | 75% | 10,393,804 | 12,240 | 0 | 0 | 0 |
| Extended-NPI | 20+ | 90% | 10,108,108 | 11,932 | 0 | 0 | 0 |
| Extended-NPI | All | 0% | 11,472,404 | 13,216 | 0 | 0 | 0 |
| Extended-NPI | All | 50% | 9,654,148 | 11,192 | 0 | 0 | 0 |
| Extended-NPI | All | 75% | 9,546,620 | 11,296 | 0 | 0 | 0 |
| Extended-NPI | All | 90% | 9,418,868 | 10,680 | 0 | 0 | 0 |
| Intensive-NPI | 20+ | 0% | 30,157,284 | 61,028 | 11,390 | 8,390 | 4,390 |
| Intensive-NPI | 20+ | 50% | 26,995,304 | 53,364 | 9,156 | 6,156 | 2,156 |
| Intensive-NPI | 20+ | 75% | 25,390,024 | 48,188 | 7,654 | 4,654 | 654 |
| Intensive-NPI | 20+ | 90% | 24,700,932 | 47,396 | 7,248 | 4,248 | 248 |
| Intensive-NPI | All | 0% | 28,473,828 | 53,704 | 8,852 | 5,852 | 1,852 |
| Intensive-NPI | All | 50% | 22,260,956 | 39,664 | 4,100 | 1,100 | 0 |
| Intensive-NPI | All | 75% | 19,502,680 | 33,992 | 2,192 | 0 | 0 |
| Intensive-NPI | All | 90% | 17,733,452 | 29,944 | 1,204 | 0 | 0 |
| $p_A = 35\%$ | | | | | | | |
| Extended-NPI | 20+ | 0% | 15,248,560 | 12,720 | 0 | 0 | 0 |
| Extended-NPI | 20+ | 50% | 10,819,820 | 10,376 | 0 | 0 | 0 |
| Extended-NPI | 20+ | 75% | 10,524,788 | 9,976 | 0 | 0 | 0 |
| Extended-NPI | 20+ | 90% | 10,166,996 | 9,756 | 0 | 0 | 0 |
| Extended-NPI | All | 0% | 12,689,328 | 12,332 | 0 | 0 | 0 |
| Extended-NPI | All | 50% | 9,716,924 | 9,208 | 0 | 0 | 0 |
| Extended-NPI | All | 75% | 9,578,812 | 9,488 | 0 | 0 | 0 |
| Extended-NPI | All | 90% | 9,464,388 | 8,920 | 0 | 0 | 0 |
| Intensive-NPI | 20+ | 0% | 30,684,416 | 51,756 | 9,506 | 6,506 | 2,506 |
| Intensive-NPI | 20+ | 50% | 27,121,000 | 43,892 | 6,702 | 3,702 | 0 |
| Intensive-NPI | 20+ | 75% | 25,391,704 | 39,256 | 5,350 | 2,350 | 0 |
| Intensive-NPI | 20+ | 90% | 24,469,140 | 37,936 | 4,710 | 1,710 | 0 |
| Intensive-NPI | All | 0% | 29,061,100 | 44,164 | 6,210 | 3,210 | 0 |
| Intensive-NPI | All | 50% | 22,642,780 | 33,556 | 2,610 | 0 | 0 |
| Intensive-NPI | All | 75% | 19,287,024 | 27,216 | 834 | 0 | 0 |
| Intensive-NPI | All | 90% | 18,370,684 | 25,968 | 634 | 0 | 0 |

Applying the Intensive-NPI, the number of deaths was three times higher than with a 6-month vaccination campaign under similar conditions.

In all simulations with Intensive-NPI, the ICU resources were saturated. The application of the extended-NPI made possible to avoid the ICU resources saturation.

With a 12-month vaccination campaign of all individuals without transmission reduction (Table 6) vs. no vaccination and Intensive-NPI (Table 4), the number of deaths was divided by 3 with the Intensive-NPI and by 10 with the Extended-NPI.

### 3.5 Effects of 18-month and 24-month vaccination campaigns

Tables 7 and 8 show the results obtained with an 18-month and a 24-month vaccination campaign respectively.

**Table 7:** Results of an 18-month vaccination campaign with the historical strain and new variants: two-years cumulative numbers of individuals removed and deaths, ICU overload exceeding beds capacity under different restrictions, different vaccination options and different transmission reduction.

| NPI | Vaccination<br>Option | Transmission<br>Reduction | Removed | Deceased | ICU overload if max available beds is |  |  |
| --- | --- | --- | --- | --- | --- | --- | --- |
|  |  |  |  |  | 5,000 | 8,000 | 12,000 |
| $p_A = 20\%$ | | | | | | | |
| Extended-NPI | 20+ | 0% | 17,694,840 | 26,796 | 0 | 0 | 0 |
| Extended-NPI | 20+ | 50% | 13,818,360 | 20,060 | 0 | 0 | 0 |
| Extended-NPI | 20+ | 75% | 12,532,892 | 17,136 | 0 | 0 | 0 |
| Extended-NPI | 20+ | 90% | 12,841,648 | 18,584 | 0 | 0 | 0 |
| Extended-NPI | All | 0% | 15,518,292 | 22,240 | 0 | 0 | 0 |
| Extended-NPI | All | 50% | 11,473,312 | 15,536 | 0 | 0 | 0 |
| Extended-NPI | All | 75% | 10,760,576 | 14,076 | 0 | 0 | 0 |
| Extended-NPI | All | 90% | 10,508,564 | 13,580 | 0 | 0 | 0 |
| Intensive-NPI | 20+ | 0% | 31,823,672 | 81,604 | 18,960 | 15,960 | 11,960 |
| Intensive-NPI | 20+ | 50% | 30,758,828 | 77,852 | 17,886 | 14,886 | 10,886 |
| Intensive-NPI | 20+ | 75% | 30,320,220 | 75,132 | 17,386 | 14,386 | 10,386 |
| Intensive-NPI | 20+ | 90% | 29,913,664 | 74,404 | 16,978 | 13,978 | 9,978 |
| Intensive-NPI | All | 0% | 30,873,760 | 74,092 | 15,546 | 12,546 | 8,546 |
| Intensive-NPI | All | 50% | 28,556,484 | 66,344 | 13,640 | 10,640 | 6,640 |
| Intensive-NPI | All | 75% | 27,355,052 | 61,976 | 12,526 | 9,526 | 5,526 |
| Intensive-NPI | All | 90% | 26,686,632 | 60,880 | 11,558 | 8,558 | 4,558 |
| $p_A = 35\%$ | | | | | | | |
| Extended-NPI | 20+ | 0% | 18,287,688 | 22,444 | 0 | 0 | 0 |
| Extended-NPI | 20+ | 50% | 14,467,432 | 17,816 | 0 | 0 | 0 |
| Extended-NPI | 20+ | 75% | 13,361,520 | 15,720 | 0 | 0 | 0 |
| Extended-NPI | 20+ | 90% | 12,407,172 | 13,940 | 0 | 0 | 0 |
| Extended-NPI | All | 0% | 15,829,472 | 18,788 | 0 | 0 | 0 |
| Extended-NPI | All | 50% | 11,959,612 | 13,228 | 0 | 0 | 0 |
| Extended-NPI | All | 75% | 10,779,828 | 11,276 | 0 | 0 | 0 |
| Extended-NPI | All | 90% | 10,659,880 | 11,428 | 0 | 0 | 0 |
| Intensive-NPI | 20+ | 0% | 31,903,712 | 66,628 | 14,052 | 11,052 | 7,052 |
| Intensive-NPI | 20+ | 50% | 30,883,968 | 63,168 | 13,258 | 10,258 | 6,258 |
| Intensive-NPI | 20+ | 75% | 30,458,448 | 62,136 | 13,750 | 10,750 | 6,750 |
| Intensive-NPI | 20+ | 90% | 30,140,116 | 61,904 | 13,644 | 10,644 | 6,644 |
| Intensive-NPI | All | 0% | 31,220,868 | 60,296 | 12,030 | 9,030 | 5,030 |
| Intensive-NPI | All | 50% | 28,770,572 | 54,780 | 10,326 | 7,326 | 3,326 |
| Intensive-NPI | All | 75% | 27,680,844 | 51,328 | 9,600 | 6,600 | 2,600 |
| Intensive-NPI | All | 90% | 26,840,120 | 49,532 | 8,804 | 5,804 | 1,804 |

**Table 8:** Results of a 24-month vaccination campaign with the historical strain and new variants: two-years cumulative numbers of individuals removed and deaths, ICU overload exceeding beds capacity under different restrictions, different vaccination options and different transmission reduction.

| NPI | Vaccination<br>Option | Transmission<br>Reduction | Removed | Deceased | ICU overload if max available beds is |  |  |
| --- | --- | --- | --- | --- | --- | --- | --- |
|  |  |  |  |  | 5,000 | 8,000 | 12,000 |
| $p_A = 20\%$ | | | | | | | |
| Extended-NPI | 20+ | 0% | 19,752,664 | 37,076 | 0 | 0 | 0 |
| Extended-NPI | 20+ | 50% | 17,295,276 | 30,628 | 0 | 0 | 0 |
| Extended-NPI | 20+ | 75% | 16,568,408 | 29,392 | 0 | 0 | 0 |
| Extended-NPI | 20+ | 90% | 15,742,112 | 27,068 | 0 | 0 | 0 |
| Extended-NPI | All | 0% | 17,984,716 | 31,264 | 0 | 0 | 0 |
| Extended-NPI | All | 50% | 14,045,300 | 22,156 | 0 | 0 | 0 |
| Extended-NPI | All | 75% | 13,021,360 | 20,116 | 0 | 0 | 0 |
| Extended-NPI | All | 90% | 12,858,040 | 19,800 | 0 | 0 | 0 |
| Intensive-NPI | 20+ | 0% | 32,241,808 | 93,300 | 21,106 | 18,106 | 14,106 |
| Intensive-NPI | 20+ | 50% | 31,961,180 | 91,888 | 20,804 | 17,804 | 13,804 |
| Intensive-NPI | 20+ | 75% | 31,827,348 | 89,828 | 20,260 | 17,260 | 13,260 |
| Intensive-NPI | 20+ | 90% | 31,699,060 | 90,332 | 19,612 | 16,612 | 12,612 |
| Intensive-NPI | All | 0% | 31,860,848 | 83,620 | 19,396 | 16,396 | 12,396 |
| Intensive-NPI | All | 50% | 30,987,580 | 80,728 | 18,458 | 15,458 | 11,458 |
| Intensive-NPI | All | 75% | 30,689,824 | 80,556 | 18,452 | 15,452 | 11,452 |
| Intensive-NPI | All | 90% | 30,371,528 | 79,900 | 18,202 | 15,202 | 11,202 |
| $p_A = 35\%$ | | | | | | | |
| Extended-NPI | 20+ | 0% | 20,012,972 | 29,968 | 0 | 0 | 0 |
| Extended-NPI | 20+ | 50% | 17,454,864 | 25,188 | 0 | 0 | 0 |
| Extended-NPI | 20+ | 75% | 16,442,256 | 23,252 | 0 | 0 | 0 |
| Extended-NPI | 20+ | 90% | 15,856,440 | 22,092 | 0 | 0 | 0 |
| Extended-NPI | All | 0% | 18,207,032 | 25,172 | 0 | 0 | 0 |
| Extended-NPI | All | 50% | 14,498,900 | 19,588 | 0 | 0 | 0 |
| Extended-NPI | All | 75% | 13,280,304 | 17,148 | 0 | 0 | 0 |
| Extended-NPI | All | 90% | 12,494,804 | 15,572 | 0 | 0 | 0 |
| Intensive-NPI | 20+ | 0% | 32,341,944 | 76,244 | 16,344 | 13,344 | 9,344 |
| Intensive-NPI | 20+ | 50% | 32,009,816 | 74,532 | 15,996 | 12,996 | 8,996 |
| Intensive-NPI | 20+ | 75% | 31,802,816 | 74,200 | 15,630 | 12,630 | 8,630 |
| Intensive-NPI | 20+ | 90% | 31,761,572 | 73,364 | 15,370 | 12,370 | 8,370 |
| Intensive-NPI | All | 0% | 32,082,144 | 69,580 | 15,108 | 12,108 | 8,108 |
| Intensive-NPI | All | 50% | 31,018,988 | 65,672 | 14,170 | 11,170 | 7,170 |
| Intensive-NPI | All | 75% | 30,714,560 | 65,228 | 14,078 | 11,078 | 7,078 |
| Intensive-NPI | All | 90% | 30,356,664 | 64,408 | 13,600 | 10,600 | 6,600 |

With the Intensive-NPI, whatever the simulated conditions, the ICU resources were saturated. The level of intensive care saturation increased markedly between the 12 and the 18 month vaccination campaigns, and continued to increase but to a lesser extent between the 18 and the 24 month vaccination campaigns.

In all vaccination campaigns exceeding 6 months, the Extended-NPI was necessary to avoid ICU resource saturation.

Versus no vaccination and Intensive-NPI (Table 4), an 18-month vaccination campaign of all individuals without transmission reduction divided the number of deaths by 2 with the Intensive-NPI and by 6 with the Extended-NPI (Table 7), whereas a 24-month vaccination campaign of all individuals without transmission reduction divided the number of deaths by 2 with the Intensive-NPI and by 4 with the Extended-NPI(Table 8).

### 3.6 Determinants of the vaccination campaign efficacy

Figure 3 shows the results of the simulations of the epidemic from January 1, 2021, depending on the duration of the vaccination campaign, the reduction in the risk of viral transmission, the prevalence of asymptomatic forms and the Intensive/Extensive type of NPI. The alternative between the Intensive-NPI / Extended-NPI strongly impacted the mortality and the ICU saturation. For a 2-year vaccination campaign restricted to individuals aged 20 or older, without reduction in viral transmission, with a prevalence of asymptomatic forms of 20%, the Intensive-NPI led to 93.3 thousand deaths while 21.1 thousand patients exceeded the ICU resources, whereas the Extended-NPI led to 37.1 thousand deaths without ICU saturation. Limiting the duration of the vaccination campaign to 6 months would lead, under the same other conditions, to 24.7 thousand deaths without ICU saturation under the Intensive-NPI, and to 11.2 thousand deaths without ICU saturation under the Extended-NPI. In case the 6-month strategy was not realistic, it was mandatory to limit the duration of the vaccination campaign and apply the Extended-NPI to limit the number of deaths and to avoid the ICU saturation.

**Figure 3:**
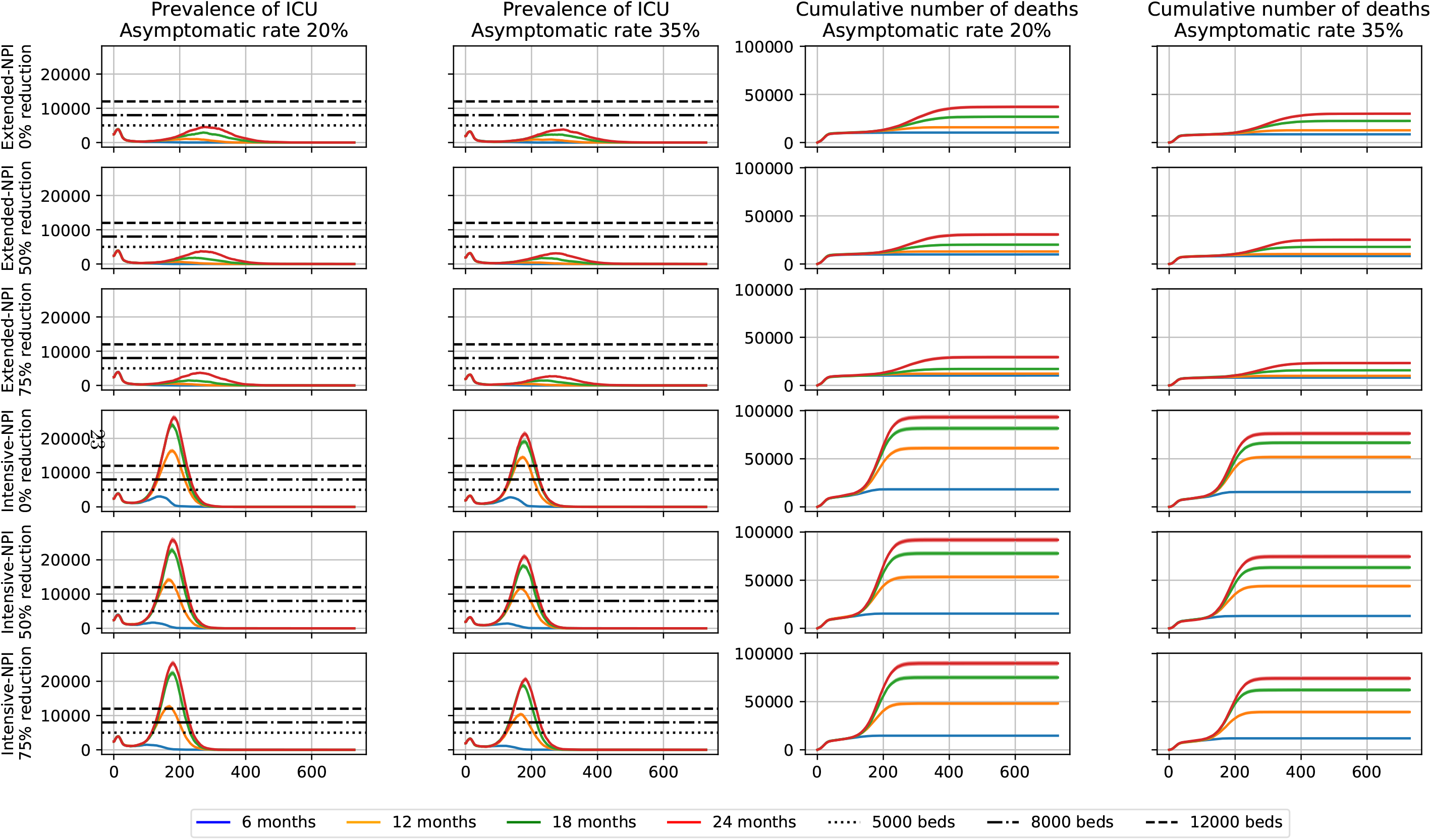
Projection of the COVID-19 pandemic in France for the historical strain and new variants, for different durations of vaccination campaigns and for reduction virus transmission of 0%, 50% and 75%. Each row displays the prevalence of COVID-19 patients in needs of ICU and the cumulative number of deaths at hospital, and for 20% and 35% asymptomatic rates.

## 4 DISCUSSION

### 4.1 Historical strain without vaccination

Table 3 presents the cumulative number of individuals removed due to disease, the cumulative number of disease-related deaths and ICU overload after two years with the historical strain and without vaccination, under Relaxed-NPI, Intensive-NPI and Extended-NPI strategies. It shows that adopting a Relaxed-NPI would lead to an important increase in the number of deaths, whatever the probability of asymptomatic form (either 20% or 35%). However, without a variant and even without a vaccine, it would not have been mandatory to switch to Extended-NPI; the Intensive-NPI would have been acceptable while waiting for a sufficient herd immunity, although the number of deaths generated would have been naturally higher than the one expected with Extended-NPI. Figure 1 shows also that Relaxed-NPI would have been sufficient to induce a virus extinction within approximately 500 days, and limit the economic cost of the sanitary situation, but at a certain cost in terms of deaths and ICU beds, and provided these NPI were continuously maintained.

### 4.2 Introduction of new variant strains without vaccination

Figure 2 shows that the Relaxed-NPI are still sufficient to lead to an extinction of both historical and variant strain virus within approximately 250 days, but at very high costs in terms of deaths (170 to 210 thousand) and of ICU beds (35 thousand patients would be deprived). With the introduction of variants, Relaxed-NPI would not be a reasonable option.

Strikingly, as soon as the variant strain is included in the computations, the Intensive-NPI works no more in limiting sufficiently the number of deaths and maintaining an acceptable pressure on ICU beds. The emergence of more infectious variant strains of the virus makes ineffective both Relaxed-NPI and Intensive-NPI. Table 4 shows that it is even necessary to strengthen Intensive-NPI and to impose Extended-NPI to avoid an explosion of the number of deaths, and a total overload of the ICU resources. The level of Herd immunity then reached does not make possible to release the NPI because it does not protect against a new epidemic wave once these protective measures have been withdrawn.

Without vaccines, 550 days would be necessary to halt the epidemic, and Extended-NPI would be inevitable. Without vaccines and with new variants, using simply current preventive measures would be very costly in terms of deaths, ICU beds, and paralysis of the health care system. It is then important to accelerate the vaccination campaigns. Increasing herd immunity through vaccination is necessary to avoid the resumption of the epidemic after NPI removal.

### 4.3 Adapting the vaccine strategy to new variant strains

In the absence of vaccination, the emergence of a new variant strain changes definitely the dynamics of the epidemic. The vaccination strategy has to be adapted to a new epidemic dynamics. The present study aims to comparing the impacts of 4 vaccination strategies to face a new epidemic dynamics (6, 12, 18 and 24-month campaigns).

The main conclusion of modelling the vaccination strategy effect is that the best ways to deal with a historical and a new strain, without overloading ICUs is to vaccinate the entire population or all persons aged over 20 within 6 months (The minimum age for vaccination will depend on the results of ongoing pediatric clinical trials). This is shown in Table 5 where a maximal of 18 thousand deaths and no ICU overload are observed with applying the Intensive-NPI. Currently, vaccinating the whole French population within 6 months does not seem to be an achievable goal; the vaccination campaign has begun on January 11, 2021, and on March 13, 2021, 2,220,608 people have been already fully vaccinated (3, 3% of the population). Thus, despite an ongoing significant acceleration, the goal would still not be achievable.

Tables 6, 7 and 8 show the potential effects of 12, 18 and 24 month vaccination campaigns. Despite intensive NPI, vaccination campaigns lasting 18 months or 24 months would generate up to 81 and 93 thousand deaths respectively, and significant pressure on ICU beds in both scenarios.

Taking into account the currently unlikely vaccination strategy within 6 months, it appears that vaccinating the whole population within 1 year would bring real benefits in terms of deaths and ICU saturation. As shown in Table 6, the 12-month vaccination strategy provides reasonable results with Extended-NPI. By contrast, with Intensive-NPI, the number of deaths may reach 61 thousand in the worst scenario and exceed 20 to 30 thousand in many others whereas the pressure on ICU beds would exceed the maximum capacities by several thousand. With the Intensive-NPI scenario, there are real risks of paralyzing the health-care system. The only solution to overcome the possible failure of a 6-month vaccination strategy seems to be the adoption of Extended-NPI and the vaccination campaign within a maximum of 12 months.

The need for Extended-NPI seems to be inevitable, at least during a transitory period. Relaxing Intensive-NPI will be definitely a crucial issue for economic and social reasons. It will highly depend on the number of vaccinated persons: i.e. as long as the number of vaccinated persons is insufficient to protect the population and to obtain herd immunity, it will be difficult to release the NPI without worsening additional deaths and ICU pressures. One notice that the situation would be far better than a vaccination strategy over more than 12 months. An important additional argument in favour of limiting the duration of the vaccination campaign is the difficulty to maintain the acceptation of the appropriate NPI at population level over a large period of time.

It is important to mention that in all extended-NPI scenarios, the duration of the vaccination campaign impacts the number of deaths: 9-10 thousand with the 6-month strategy, 11-16 thousand with the 12 months strategy, 14-27 thousand with the 18 month strategy and 20-37 thousand with the 24-month one. Although ICU resources are not overwhelmed applying the extended-NPI, the duration of the vaccination campaign strongly influences their use (Figure 3). The hospitalization of COVID-19 patients in intensive care significantly disrupts medical activity, forcing the deprogramming of much of usual medical and surgical activity. There is a real need to accelerate the vaccination of “the whole population in one year” to maintain care capacities, treat patients in intensive care and avoid excessive deaths.

The present study makes the strong hypothesis of a 100% acceptation level of vaccination. Virus extinction will happen if the population is widely vaccinated. This means that more efforts should be made in terms of vaccination strategy, and wise communication regarding the vaccines. It would be interesting to tackle the issue of vaccine acceptation and measure the impacts of partial acceptation levels on the extinction of the pandemic.The adverse effects of various vaccines do influence vaccination strategies and the vaccine acceptation level. The present modeling deliberately ignored the side effects of COVID-19 vaccines, these being considered negligible compared to the expected benefits of establishing herd vaccine immunity. Another choice the modelling was ignoring the duration of vaccine protection. This might be the object of future investigations

## 5 Conclusion

The main conclusion of this study is that the key point of this race against the COVID-19 historical strain and its variant strains is an issue of vaccination strategy. It is mandatory to vaccinate most of the population within a year, and preferably within 6 months. Should a 6 month vaccination campaign not be feasible, then reinforced NPI should be considered.

These conclusions are in agreement with the recommendations of Bosetti et al. [18] that reinforced NPI applied to the most contagious variant are necessary to achieve epidemic control similar to that obtained by applying less stringent measures to the historical virus strain. These authors also insist that keeping transmission rates of the virus low helps prevent a worsening of the epidemic requiring enhanced NPI.

One advantage of the modeling presented in this paper is that it takes into account the particularity of the French vaccine strategy: vaccinating the most advanced age groups first. If it appears that it will not be possible to vaccinate all the population within one year, alternative solutions will have to be examined. Regarding accelerating vaccination strategy would it be more impactful to increase the delay between the two doses or increase the pace of vaccination through increasing the numbers of vaccination centers, vaccination human resources or available vaccine supplies? Answering this question is not the aim of the present report. Nevertheless, it seems important to mention that solutions such as increasing the delay between the two doses of vaccines could have a direct impact on the success of the vaccination, on its efficiency.

Another advantage of rapid vaccination is to help reduce the worldwide multiplication of the virus, and thus to limit the possible emergence of vaccine-resistant variants, and/or more virulent ones. The present paper is only calibrated on the historical strain and on the variants with contagiousness similar to the SARS-CoV-2VOC 202012/01 (B.1.1.7), a virulence similar to the historic strain, and without specific vaccine-resistance. The available data don’t allow to examine the effect of the emergence of more strains. Nevertheless, as soon as enough data are available to estimate a realistic *β*_3_ for a new variant, the present model is a useful tool to compare the effects of various vaccine strategies on new variant strains. Studying other variants requires measuring the efficiency of the vaccines on these variants. It is possible to integrate a vaccine-resistance parameter specific to the variant in the model (variants such as B.1.351 [17]), as soon as studies on the subject could provide serious estimations of such parameter. An increase of the virulence of the virus with a variant strain could be treated by modifying the transition probabilities in the model (see [19]).

## Data Availability

All data used during this study are available on data.gouv.fr or in the published articles listed in the references, except APHP related data obtained from a personal communication of the EHESP. All data generated during this study are available from the corresponding author on reasonable request.

## Members of the CovDyn (Covid Dynamics) group, by main affiliation

**LBBE – HCL** Mathieu FAUVERNIER, François GUEYFFIER, Jean IWAZ, Delphine MAUCORT-BOULCH, Simon PAGEAUD, Muriel RABILLOUD, Pascal ROY.

**LSAF** Alexis BIENVENÜ E, Anne EYRAUD-LOISEL, Romain GAUCHON, Pierre-Olivier GOFFARD, Nicolas LEBOISNE, Stéphane LOISEL, Xavier MILHAUD, Denys POMMERET, Yahia SALHI.

**LAMA** Pierre VANDEKERKHOVE.

**CIRI-HCL** Philippe VANHEMS.

**LMFA** Jean-Pierre BERTOGLIO.

**LTDS** Nicolas PONTHUS.

**INL** Christelle YEROMONAHOS.

**LIRIS** Stéphane DERRODE, Véronique DESLANDRES, Mohand-Saïd HACID, Salima HASSAS, Catherine POTHIER, Christophe RIGOTTI.

**ICJ** Thibault ESPINASSE, Samuel BERNARD, Philippe MICHEL, Vitaly VOLPERT.

**INRIA** Mostafa ADIMY.

**CHU De Rouen Unité INSERM 1018, CESP** Jacques BENICHOU.

**CHU De Rouen LIMICS INSERM U1142, Université de Rouen/ Sorbonne Université** Stéfan DARMONI.

**CHU de NICE, Université de Nice** Pascal STACCINI.

## Footnotes

1 Personal communication from the EHESP.

